# Life-course social participation and physical activity in midlife: Longitudinal associations in the 1970 British Cohort Study (BCS70)

**DOI:** 10.1101/2022.10.21.22281357

**Authors:** Stergiani Tsoli, Daisy Fancourt, Alice Sullivan, Mark Hamer, George B. Ploubidis, Ichiro Kawachi

## Abstract

**Background:** A hypothesized benefit of social participation is that it encourages people to be more physically active. However, limited evidence exists on the association between social participation over the life-course and physical activity in midlife. We sought to apply a life-course framework to examine the association of social participation and device measured physical activity in midlife in the UK.

**Methods:** We used the 1970 British Birth Cohort Study (BCS70), which includes all people born in Britain during a single week in 1970. Social participation was assessed at ages 16, 30, 34 and 42. Physical activity was measured by accelerometery at age 46, as mean daily step count and time spent in Moderate to Vigorous Physical Activity (MVPA). The associations of social participation and physical activity were tested using two different life-course models: the sensitive period model and the accumulation model.

**Results:** Individuals with medium and high participation compared to no social participation over their life-course had higher mean daily step count and MVPA in midlife, supporting the accumulation model. In the sensitive period model, only those that actively participated at age 42 had higher mean daily steps and MVPA compared to those who did not participate.

**Conclusion:** Our study provides empirical evidence on the importance of sustaining social participation at all ages over the life-course rather than at a particular timepoint of someone’s life. Interventions to promote social participation throughout the life-course could be an avenue to promote physical activity in middle life.

## 1 Introduction

Social participation is frequently described as a person’s involvement in activities that provide interaction with others in society or the community (1). An abundance of evidence links active participation in social and leisure activities with physical and mental health outcomes in later life (2-4), including health and happiness (5), health functioning (6), age-related physical decline including disability, and frailty (7, 8), malaise (9), anxiety and depression (7, 10), quality of life (11), cognitive function (12), limiting long-standing illness (9) and health-related behaviours like alcohol use (9). A recent review (13) has suggested examples through which social participation may act as a mechanism of action for improving mental and physical health outcomes. Proposed mechanisms through which social participation promotes health and wellbeing include increased opportunities to exchange social support with others in a group, maintenance of cognitive skills (“use it or lose it”) through social interactions, and peer influence (e.g., encouragement and support to give up smoking from one’s social group).

Notably, there is also growing evidence suggesting that social participation may be linked to physical activity (14), though few studies looked at engagement in social activities and improvements in physical activity with a focus in later life (15-18). Most studies have been cross-sectional using self-report to measure physical activity, which could lead to reverse causation bias.

The way a life unfolds and people transit through different life stages can lead to accumulation or loss of the resources related with social engagement (2). Adopting a life-course approach helps us to understand the dynamic relationship between social participation and physical activity in later life. For example, a latency model would posit that physical activity during social participation in adolescence (e.g., through membership in a youth organization) confers a lifelong habit of being active, even if the individual drops out of participating in social groups throughout early adulthood and midlife because of work and family obligations. Furthermore, adolescence may be a developmentally sensitive period during which lifelong habits of physical activity are acquired through social participation. In contrast, an accumulation approach proposes that lifelong persistent social participation is important for effects to accumulate such that they might influence physical activity later in life. Previous research has examined associations of social participation and physical activity primarily at specific age periods (15, 16) and more commonly in social participation midlife or later. However, none of the previous work mentioned tested the two models.

Therefore, in a UK general population sample, we aimed to identify the association of social participation, as a potential indicator of social capital and objectively measured physical activity in midlife, using a life-course framework. We examined (i) the independent association of social participation at different periods through the life-course on physical activity at age 46 and, (ii) the life-course associations of cumulative social participation between the ages of 16-42 on physical activity at age 46 in BCS70. Possible life-course pathways to adult physical activity are shown in Figure 1.

**Figure 1:**
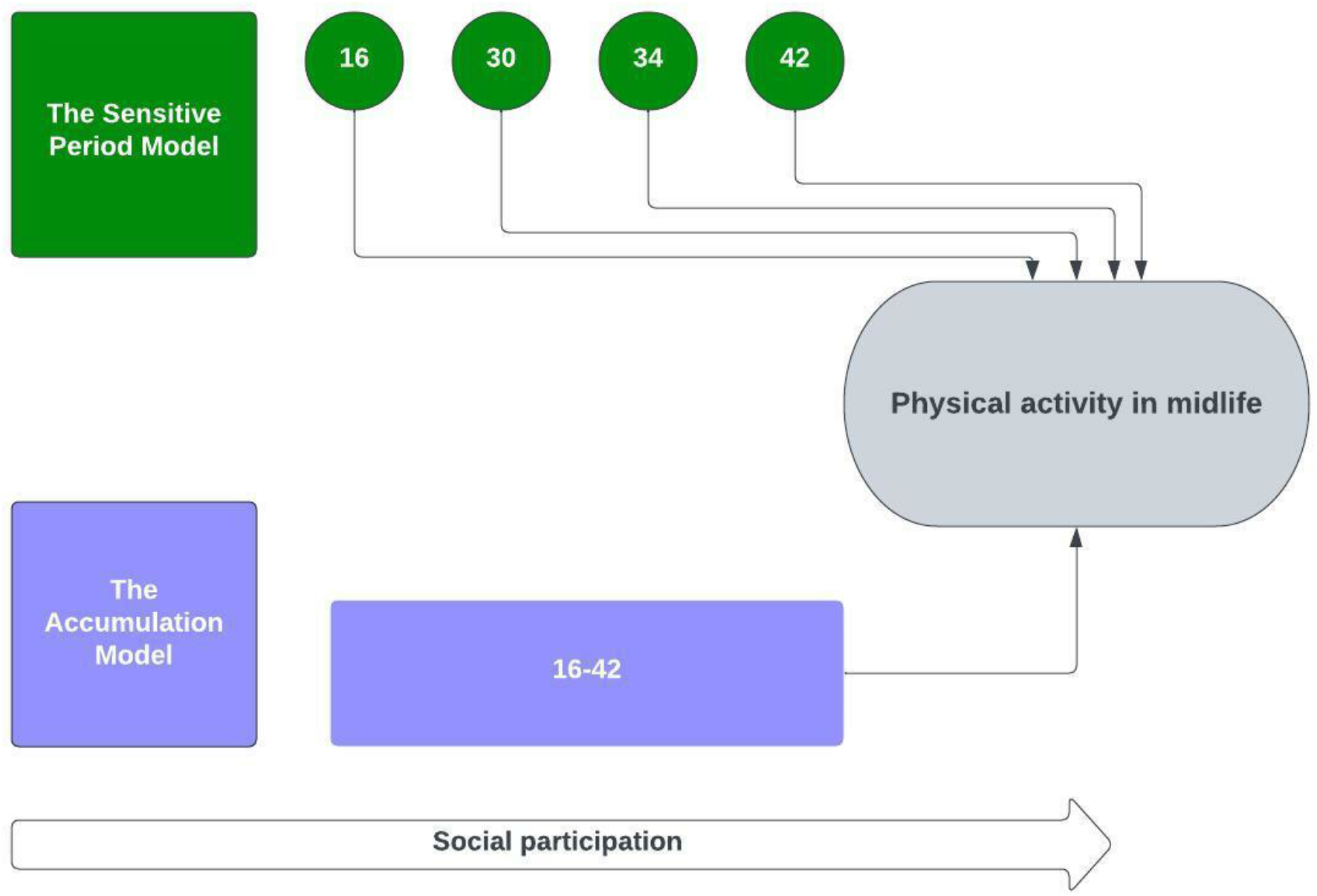
Hypothesized relationships between social participation during the life-course and mid-life physical activity.

## 2 Materials and Methods

### 2.1 Participants

We used data from the 1970 British Cohort Study (BCS70), a national birth cohort study that follows the lives of 17,196 individuals born in England, Scotland, and Wales in a single week in April 1970 (19-21). Among other factors, BCS70 collects information on participants’ health, educational, social, and physical development. The participants have been followed-up nine times since the first survey between the ages 5 and 46. The study remains representative of the original sample despite attrition (22).

Our analytical sample includes all participants in BCS70 at Wave 10 at age 46 (2016) with 8,581 participants. From the 8,581 participants at the Age 46 Survey, 6,492 provided consent to wear the monitor and 5,569 provided data for at least one day. In our analysis, we only considered those that had valid measurements of objectively measured physical activity for a week (as per the study’s protocol), which resulted in an analytical sample of 3,646 participants (see Figure 2). Those who died or emigrated by age 46 were excluded from our analytical sample. Ethical approval was received for BCS70 from the NHS Multi-Centre Research Ethics Committee (MREC) with informed consent given by the participants.

**Figure 2:**
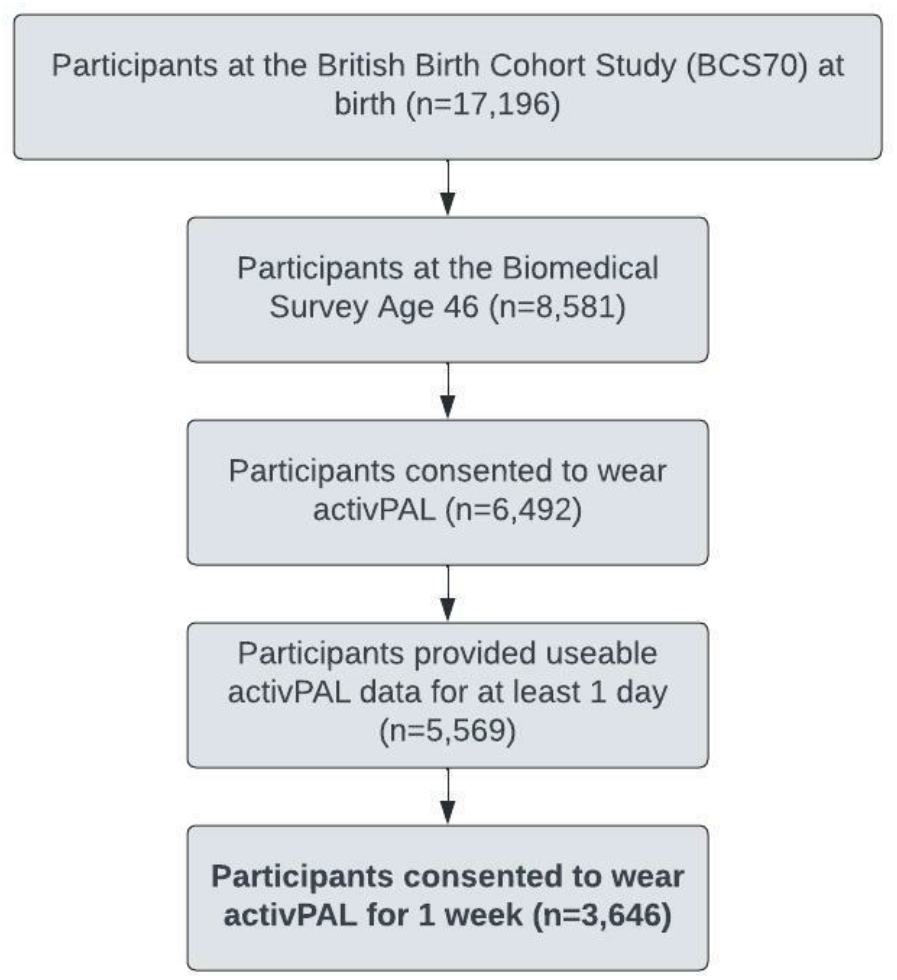
Sample selection for this study.

### 2.2 Measures

#### 2.2.1 Social participation

Information on social participation was assessed at ages 16, 30, 34, and 42. At age 16 the participants were asked whether they belong to any uniformed youth organisations. At later sweeps they were asked whether they engaged with a number of activities including political parties, trade unions, environmental groups, tenants/ residents’ associations, neighbourhood watch, church or religious groups, charitable associations, evening classes, social clubs, sports clubs, or other clubs/societies (see Supplemental Table 2). We created binary indicators of participation or no participation to those activities in each sweep.

#### 2.2.2 Objective measures of physical activity

As part of the age 46 Survey, extensive biomedical measures including objective measures of physical activity were collected by a research nurse for the first time since childhood.

Cohort members were asked to wear a thigh-mounted accelerometer (the activPAL3 micro device, PAL Technologies Ltd., Glasgow, UK) (23) continuously over the course of a 7-day including during bathing, sleeping, and all physical activity. Physical activity was sampled at the default frequency of 20Hz. The feasibility of deploying this technology in a population cohort has been documented (24). A previously described wear protocol was utilised (25). The waterproofed device was fitted by a nurse on the midline anterior aspect of their upper thigh. Participants also completed a daily diary recording their sleep and wake time along with details on any removal of the device. Participants measurements were eligible if they recorded at least 10 hours of valid wear time over a day. They were advised to wear the accelerometer for 7 days.

Physical activity in our study was assessed through two main measurements: Mean daily step count and Moderate to Vigorous Physical Activity (MVPA) (hr/day). MVPA was derived using a step cadence threshold ≥ 100 (26). Additional information about the protocol implemented and the measurements can be found in the Accelerometery User Guide (27).

#### 2.2.3 Potential confounders

We included a range of demographic, socioeconomic and health related factors. These are factors potentially associated with the exposure and the outcome that were not on the causal pathway between these variables. These included breastfeeding, mother’s marital status (birth), if the mother was a teen during pregnancy, parental education (birth), parental employment (birth), father’s social class (at birth and age 16), household tenure (age 5), overcrowding (>1 person per room at age 5), access to house amenities (age 5 and 16), number of family moves (age 5), parents reading weekly to child (age 5).

For ages 30-42 cohort members we included self-reported measures of social class, highest level of educational qualification, marital status, and employment.

Health-relates confounders included birth weight, smoking during pregnancy, maternal mental health (age 5), bed wetting (age 5), cognitive ability (age 10), health conditions (age 10), hospital admissions (age 10), Body Mass Index (BMI) (age 10, 16, 30, 34, 42), internalising and externalising problems (age 16), physical ability (age 16) and mental health morbidity (age 16, 30, 34, 42). Finally, we included a measure of long-standing/limiting illness, disability of infirmity and self-rated health for ages 30, 34 and 42.

A detailed description of the potential confounders, exposure and outcome variables is available in Supplemental Table 2.

## 3 Statistical analysis

In our main analysis, we examined the relationship of between social participation and physical activity outcomes measured at age 44-46 as continuous variables using ordinary least squares (OLS) regressions models. We evaluated 2 life-course frameworks through 2 models (see Figure 1):

In **Model 1 (*Accumulation model*)**, we examined the association of a cumulative social participation through the life-course from age 16-42 and physical activity at midlife. A Cumulative Index of social participation (Low, Medium and High) was derived using responses at age 16, 30, 34 and 42. Social participation was coded using “None” as the reference category if cohorts members had a negative response in all four sweeps(=0), “Low” if they engaged with activities one sweep(=1), “Medium” if they engaged 2 times(=2) and “High” if engaged at least three times(≥3), respectively.

In **Model 2 (*Sensitive period model*)**, we examined the association of social participation at 4 different potentially sensitive windows of time (age 16, 30, 24 and 42) and physical activity in midlife.

We found no sex differences or interaction effects thus our analysis combined men and women.

To check the robustness of our findings, we conducted three sensitivity analyses: (i) we repeated the analysis for Model 1 (accumulation model) and restricted the responders to different scenarios: a. those with one day of valid accelerometer readings, b. those that participated at the Biomedical survey and the whole c. sample removing only those that migrated and are dead at age 46 (see Supplemental Table 4). Since the questions were not identical in all sweeps:

(ii) At age 42, cohort members were asked whether they engaged with several activities including sports clubs. Given the high correlation this specific type of social participation has with physical activity, we repeated the analysis by omitting it from the social participation indicator (see Supplemental Table 6).

(iii) In all adult waves, when cohort members were asked about their social participation, they were offered a list of suggested activities or organisations and an open category as “Other” that they could have participated. At age 30 the “other” option was not available as a category. Thus contribution of age 30 could be potentially not equivalent to the other ages (see Supplemental Table 7). So we repeated the analysis, omitting age 30 from the models.

### 3.1 Missing Data Strategy

We attempted to restore sample representativeness, increase power, and reduce bias in out estimations by employing multiple imputation with chained equations (MICE) (28-30) and generated 50 imputed datasets. Information on attrition and non-response on BCS70 has been documented elsewhere (31). More information on the missing data strategy applied in our analysis can be found in (Supplemental Text 1).

All analyses were carried out using Stata 17. 0 (32).

## 4 Results

Participants reported a mean (SD) of 0.9 (0.4) hours of daily MVPA and 9570.2 (3482) of mean daily steps. A total of 49.2% reported participating in uniformed organisations at age 16, 30.8% at age 30, 52.3% at age 34 and 60.5% for age 42, respectively.

Descriptive statistics for covariates for our sample are shown in Supplemental Table 1.

The results of the adjusted analyses of social participation and physical activity evaluated by the two life-course models are presented in **Error! Reference source not found**.. Crude estimates are, also, available (see Supplemental Table 4).

### 4.1 Accumulation Model

Evaluating the association between accumulated social participation between the age of 16 and 42 indicated higher daily step count for those in the medium and high category compared to those with no social participation at any life stage. A positive association was observed for all participation index categories and hours per day engaging in MVPA. The effect was more pronounced to those with Medium and High participation over their life-course compared to those in the Low category (see Figure 3).

**Figure 3:**
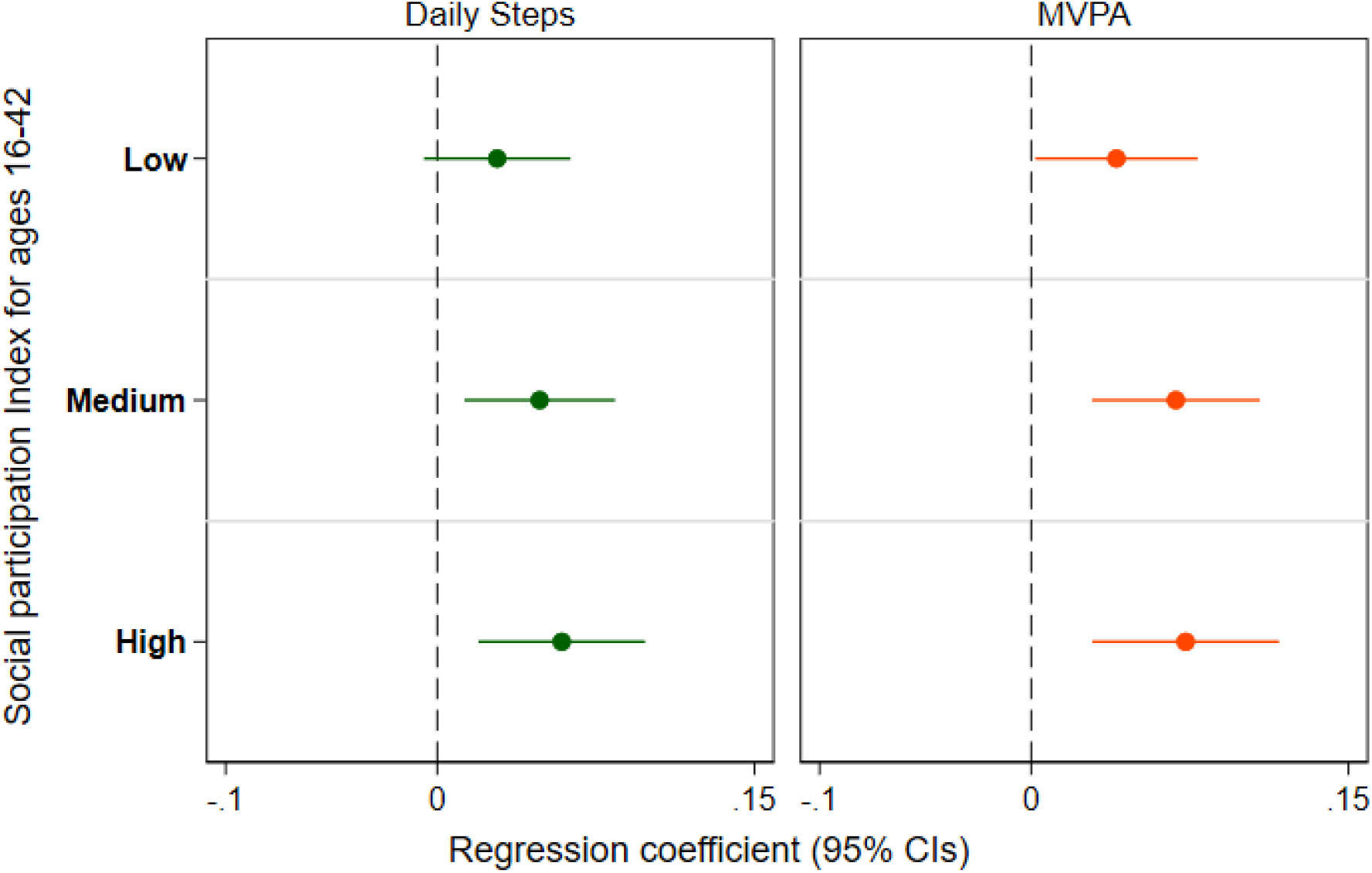
Adjusted regression coefficients (95% CI) for the a. Accumulation Model by each physical activity measure. (Reference category: No social participation from ages 16-42)

### 4.2 Sensitive period Model

For the sensitive period model, we found no evidence of association for social participation for most of the potentially sensitive windows and daily steps or MVPA. However, later life social participation was associated with an increase in both physical activity outcomes (see Table 1. And Supplemental Figure 1.) This leads us to reject the hypothesis that adolescence is a sensitive developmental period for the acquisition of habits of physical activity via social participation. It suggests that the strongest association is between recent social participation and current physical activity.

**Table 1:** Adjusted regression coefficients (95% CIs) estimating the association between social participation and physical activity at age 46 (n = 3,646) for a. Accumulation and b. Sensitive Period Model.

|  | Mean daily step count<br>(n=3,646) | Moderate to Vigorous Physical Activity (MVPA)<br>(hr/day)<br>(n=3,646) |
| --- | --- | --- |
| **a. Accumulation Model: Social Participation Index<sup>a</sup>** |  |  |
| Low | 0.028(-0.006 - 0.063) | 0.040(0.002 - 0.079)** |
| Medium | 0.048(0.013 - 0.084)*** | 0.068(0.029 - 0.108)*** |
| High | 0.059(0.019 - 0.098)*** | 0.073(0.029 - 0.117)*** |
| **b. Sensitive Period Model** |  |  |
| Age 16<sup>a</sup> | -0.012(-0.045 - 0.021) | -0.013(-0.049 - 0.022) |
| Age 30<sup>b</sup> | 0.020(-0.007 - 0.047) | 0.025(-0.006 - 0.056) |
| Age 34<sup>c</sup> | 0.012(-0.014 - 0.039) | 0.013(-0.018 - 0.043) |
| Age 42<sup>d</sup> | 0.060(0.033 - 0.088)*** | 0.066(0.035 - 0.098)*** |

\*\*\* p<0.01, \*\* p<0.05, \* p<0.1
Notes: "None" was the reference category if cohort members had a negative response in all four sweeps (=0), "Low" if they engaged with activities only one time point (=1), "Medium" 2 times (=2) and "High" three times or more (≥3)
<sup>a</sup>Age 16: Adjusted for confounders until age 16
<sup>b</sup>Age 30: Adjusted for Age 16 + confounders at age 30
<sup>c</sup>Age 34: Adjusted for Age 30 + confounders at age 34
<sup>d</sup>Age 42: Adjusted for Age 34 + confounders at age 42

### 4.3 Sensitivity analyses

When repeating the analyses with different sample restrictions (e.g. participants with at least 1 day of accelerometer data instead of 1 week) the associations became more pronounced due to increase in power. For example, those with “Low” accumulated social participation had statistically significant increased mean daily steps. (*b* (95% CIs): 0.031 (0.001-0.061, p=0.042) (see Supplemental Table 5).

After excluding those who participated at a spots club from the social participation indicator at age 42, the results largely remained the same for the sensitive period model with small changes for social participation at age 42. For the accumulation model, the results were less pronounced compared to the main model for those in the “Medium” and “High” category. The association between “Low” participation and MVPA was no longer statistically significant (see Supplemental Table 6).

After removing social participation at age 30 (see Supplemental Table 7) from the models, the results remain broadly the same with small attenuations in the effect sizes. For the accumulation model, like the previous sensitivity analysis, the results were now less pronounced especially for those in the “Medium” and “High” category. The differences were more pronounced for the findings for MVPA.

## 5 Discussion

This study adopted a life-course approach to study the association between social engagement and objectively measured physical activity in a prospective sample of middle-aged women and men. Overall, we found that higher social engagement throughout life was associated with physical activity at middle age. The association was more pronounced for MVPA and was maintained even after controlling for a wide range of potential confounders. We found no suggestion of a latency effect, whereby social participation in adolescence is a developmentally sensitive period for being physically active in midlife. Rather, the strongest association was found between recent social participation (at age 42) and physical activity in midlife (at age 46).

While the exact mechanisms behind these beneficial associations are not fully understood, we hypothesize possible mechanisms to explain how social participation could lead to changes in physical activity. Since there were no online options for participation, people need to leave their home to participate in any of the activities e.g move to the specific place that the activity takes place and thus increasing their “non-exercise” lifestyle. A direct link between being an active participant in activities (e.g sports clubs) and the increase of physical activity is possible as another mechanism of action.

Another indirect mechanism is linked to the social capital literature and relates with the norms or the exchange of resources (such as information between the members of a specific social network) which can promote certain behaviours (14, 33) such as physical activity and facilitate knowledge sharing (34).

Our study has several strengths. To the best of our knowledge, this is the first longitudinal examination of the association of life-course social engagement and physical activity using one of the British birth cohorts. Given that increased physical activity is evidently linked to chronic disease prevention (35), risk reduction and maintaining and improving functional capacity (36) in older adults, our findings are of great importance in promoting “healthy aging” (37). We used a sample of a well-established, large, nationally representative birth cohort study (20). We capitalised on the longitudinal structure of BCS70 and included rich information on cohort member’s socioeconomic and health controls over their life-course. Furthermore, the use of high-worn accelerometers to measure physical activity rather than self-report is a gold standard approach and is one of the most efficient ways to validate physical activity in the epidemiological literature (38, 39).

It should be noted that the way we operationalised social engagement is agnostic with respect to the exact type of activities with which the cohort member engaged. The different types of activities (e.g. volunteering, union membership) may have a differential effect on physical activity outcomes (6). Furthermore, whilst we used available information from all sweeps on social engagement there is a possible misclassification due to the inconsistency of the question at age 30 and 42. For the sensitive period model, this could mean that social engagement at age 30 is not the same as the other sweeps. We could only test if membership in sports club as part of social engagement at age 42 explains the findings for the accumulation model. Future research will need to determine whether sports clubs have different patterns of accumulation or sensitive period models in relation to physical activity as due to data constraints we could not explore this specific membership in the other sweeps.

Nonetheless, we conducted sensitivity analysis to explore the robustness of our findings. Furthermore, due to data constraints we were not able to examine intensity of engagement at each sweep. A common limitation of all prospective longitudinal studies is selective attrition and the losses related with missing data. Hence, to mitigate potential bias we used multiple imputation which is an approach that capitalises on the rich observed information on BCS70 (40, 41). Objective measures of physical activity were only available at one time point during adulthood and thus we cannot generalise the findings in other age groups and may not be applicable to younger cohorts. Finally, as in all analyses with observational data, bias due to unmeasured confounding cannot be ruled out.

## 6 Conclusion

Our study expands our understanding and provides new evidence on the link between social participation and objective accelerometery-based measures of physical activity among middle-aged adults in the UK. Our study provides empirical evidence on the importance of sustaining social participation at all ages over the life-course rather than a particular timepoint of someone’s life.

Looking ahead, it is particularly important to consider the particular relevance that our findings have for schemes as social prescribing (42). Considering that physical inactivity is a leading risk factor linked to increased morbidity and mortality (43), encouraging sustained social engagement at all ages through schemes like social prescribing will be key to enhancing physical activity across the life-course.

## Supporting information

All Supplemental Tables and Figures

STROBE checklist

## Data Availability

All data used in this study are open access and can be accessed through the UKDS https://beta.ukdataservice.ac.uk/datacatalogue/series/series?id=200001

## Authors contributions

All authors contributed to the study conception and design. Material preparation and analysis were performed by Stergiani Tsoli. The first draft of the manuscript was written by Stergiani Tsoli, and all authors commented on previous versions of the manuscript. All authors read and approved the final manuscript.

## Acknowledgements

Stergiani Tsoli received funding from the Economic and Social Research Council (ESRC) +3 for her doctoral studies.

George Ploubidis received funding from ESRC [ES/W013142/1].

Daisy Fancourt received funding from the Wellcome Trust [205407/Z/16/Z] and the ESRC project WELLCOMM [ES/T006994/1].

Mark Hamer received funding from the British Heart Foundation [SP/15/6/31397] that supported the accelerometery data collection for this work.

