## Supplementary material for "Life-course social participation and physical activity in midlife: Longitudinal associations in the 1970 British Cohort Study (BCS70)": All Supplemental Tables and Figures

### Supplemental Files

#### Supplemental Table 1: Descriptive statistics of the full and analytical sample

| | Full Sample | | Analytical sample | |
|---|---|---|---|---|
| | **Frequency/ Mean** | **Percentage/ SD** | **Frequency/ Mean** | **Percentage/ SD** |
| Sex of the baby at birth | | | | |
| Male | 8906 | 51.8% | 1567 | 46.2% |
| Female | 8279 | 48.2% | 1823 | 53.8% |
| Mother's age at birth | | | | |
| Teenage pregnancy | 1666 | 9.7% | 254 | 7.5% |
| Non-teen pregnancy | 15495 | 90.3% | 3133 | 92.5% |
| Mother's marital status at birth | | | | |
| Used to be married/single | 1265 | 7.4% | 165 | 4.9% |
| Married | 15914 | 92.6% | 3221 | 95.1% |
| Mother's age at completion of education | | | | |
| Left school before minimum leaving age | 1337 | 7.9% | 206 | 6.1% |
| Stayed at school | 15680 | 92.1% | 3158 | 93.9% |
| Fathers age at completion of education | | | | |
| Left school before minimum leaving age | 1550 | 9.6% | 258 | 7.9% |
| Stayed at school | 14663 | 90.4% | 2999 | 92.1% |
| Employment status of mother at birth | | | | |
| Unemployed | 11531 | 94.1% | 2303 | 94.5% |
| Employed | 722 | 5.9% | 135 | 5.5% |
| Employment status of father at birth | | | | |
| Unemployed | 607 | 3.8% | 88 | 2.8% |
| Employed | 15218 | 96.2% | 3090 | 97.2% |
| Smoking during pregnancy | | | | |
| Never/used to smoke | 10024 | 58.6% | 2131 | 63.3% |
| Smoking during pregnancy | 7085 | 41.4% | 1237 | 36.7% |
| Birthweight (grams) | 3268.5 | 581.3 | 3321.7 | 524.5 |
| Father's social class at birth | | | | |
| i | 820 | 5.2% | 233 | 7.4% |
| ii | 1906 | 12.1% | 452 | 14.3% |
| iii non-manual | 1924 | 12.2% | 463 | 14.6% |
| iii manual | 7544 | 47.8% | 1475 | 46.6% |
| iv partly skilled | 2473 | 15.7% | 406 | 12.8% |
| v | 1106 | 7.0% | 139 | 4.4% |
| Breast Feeding | | | | |
| Never Breastfed | 8182 | 62.9% | 1753 | 58.2% |
| Breastfed | 4824 | 37.1% | 1261 | 41.8% |
| Tenure of accommodation age 5 | | | | |
| Other | 5708 | 43.6% | 1057 | 34.9% |
| Owner | 7386 | 56.4% | 1974 | 65.1% |
| Sole use of amenities in the household age 5 | | | | |
| Shared use of amenities | 612 | 4.8% | 96 | 3.3% |
| Sole use of amenities | 12087 | 95.2% | 2853 | 96.7% |
| Crowdedness in the household age 5 | | | | |

| | | | | |
|---|---|---|---|---|
| < than 2 | 12757 | 98.6% | 2971 | 99.1% |
| 2 or more | 186 | 1.4% | 27 | 0.9% |
| Family moves age 5 | | | | |
| Never | 5588 | 43.1% | 1391 | 46.2% |
| 1-2 times | 6007 | 46.3% | 1351 | 44.9% |
| 3+ times | 1383 | 10.7% | 269 | 8.9% |
| Daytime bed wetting age 5 | | | | |
| No | 11731 | 89.9% | 2727 | 90.1% |
| Yes | 1317 | 10.1% | 299 | 9.9% |
| Mother's malaise age 5 | | | | |
| High malaise 8+ | 2346 | 18.2% | 441 | 14.7% |
| Low malaise 0-7 | 10532 | 81.8% | 2556 | 85.3% |
| Medical conditions of CM age 10 | | | | |
| < than 3 | 11698 | 97.5% | 2838 | 97.7% |
| 3 or more | 300 | 2.5% | 67 | 2.3% |
| General cognitive ability at age 10 | 0.0 | 1.6 | 0.4 | 1.5 |
| No of hospital admissions age 10 | 1.3 | 0.7 | 1.3 | 0.6 |
| BMI age 10 | 16.9 | 2.1 | 16.8 | 2.0 |
| Use of facilities age 16 | | | | |
| Shared use of amenities | 1391 | 15.0% | 356 | 14.5% |
| Sole use of amenities | 7863 | 85.0% | 2101 | 85.5% |
| Father's social class age 16 | | | | |
| i | 511 | 8.0% | 170 | 8.8% |
| ii | 1888 | 29.7% | 613 | 31.9% |
| iii non-manual | 653 | 10.3% | 214 | 11.1% |
| iii manual | 2567 | 40.3% | 718 | 37.4% |
| iv partly skilled | 594 | 9.3% | 166 | 8.6% |
| v | 153 | 2.4% | 41 | 2.1% |
| Mother's marital status age 16 | | | | |
| Not living with natural parents | 2206 | 24.3% | 510 | 21.2% |
| Living with natural parents | 6855 | 75.7% | 1897 | 78.8% |
| Internalising behaviour age 16 | 2.4 | 2.2 | 2.3 | 2.1 |
| Externalising behaviour age 16 | 2.0 | 2.7 | 1.7 | 2.4 |
| Impairment/disability/handicap age 16 | | | | |
| No | 8870 | 95.2% | 2372 | 96.7% |
| Yes | 447 | 4.8% | 82 | 3.3% |
| BMI age 16 | 21.1 | 3.1 | 20.9 | 2.8 |
| Social class at age 30 | | | | |
| i | 573 | 6.3% | 219 | 7.8% |
| ii | 3168 | 34.7% | 1034 | 36.8% |
| iii non-manual | 2253 | 24.7% | 732 | 26.0% |
| iii manual | 1886 | 20.7% | 503 | 17.9% |
| iv partly skilled | 1001 | 11.0% | 265 | 9.4% |
| v | 251 | 2.7% | 60 | 2.1% |
| Marital status age 30 | | | | |
| used to be married/single | 6345 | 56.6% | 1761 | 54.0% |
| married | 4874 | 43.4% | 1502 | 46.0% |
| Main activity age 30 | | | | |
| Unemployed | 2077 | 18.5% | 443 | 13.6% |
| Employed | 9142 | 81.5% | 2820 | 86.4% |
| Long standing illness/disability age 30 | | | | |
| No | 8605 | 76.8% | 2552 | 78.2% |
| Yes | 2603 | 23.2% | 711 | 21.8% |
| General health age 30 | | | | |
| Poor | 241 | 2.1% | 36 | 1.1% |

| | | | | |
|---|---|---|---|---|
| Fair | 1436 | 12.8% | 344 | 10.5% |
| Good | 5956 | 53.1% | 1714 | 52.5% |
| Excellent | 3578 | 31.9% | 1169 | 35.8% |
| Education level age 30 | | | | |
| None | 2966 | 26.5% | 693 | 21.3% |
| Level 1 and below | 973 | 8.7% | 231 | 7.1% |
| O-levels | 3501 | 31.2% | 988 | 30.3% |
| A-levels | 737 | 6.6% | 230 | 7.1% |
| Degree/ higher degree | 3034 | 27.1% | 1116 | 34.3% |
| Malaise score at age 30 | | | | |
| High malaise 8+ | 1409 | 12.7% | 306 | 9.4% |
| Low malaise 0-7 | 9700 | 87.3% | 2937 | 90.6% |
| BMI age 30 | 24.9 | 4.3 | 24.4 | 3.9 |
| Social class at age 34 | | | | |
| i | 542 | 6.8% | 213 | 8.0% |
| ii | 3174 | 39.7% | 1140 | 42.9% |
| iii non-manual | 1664 | 20.8% | 562 | 21.2% |
| iii manual | 1518 | 19.0% | 434 | 16.3% |
| iv partly skilled | 877 | 11.0% | 260 | 9.8% |
| v | 214 | 2.7% | 48 | 1.8% |
| Marital status age 34 | | | | |
| Used to be married/single | 2455 | 25.5% | 683 | 22.0% |
| Married/cohabiting | 7185 | 74.5% | 2424 | 78.0% |
| Main activity age 34 | | | | |
| Unemployed | 1623 | 16.8% | 444 | 14.3% |
| Employed | 8013 | 83.2% | 2663 | 85.7% |
| Long standing illness/disability age 34 | | | | |
| No | 6930 | 71.8% | 2313 | 74.4% |
| Yes | 2722 | 28.2% | 794 | 25.6% |
| General health age 34 | | | | |
| Poor | 597 | 6.2% | 139 | 4.5% |
| Fair | 1426 | 14.8% | 406 | 13.1% |
| Good | 4466 | 46.4% | 1436 | 46.2% |
| Excellent | 3143 | 32.6% | 1127 | 36.3% |
| Malaise score at age 34 | | | | |
| High malaise 4+ | 1483 | 15.5% | 376 | 12.1% |
| Low malaise 0-3 | 8113 | 84.5% | 2723 | 87.9% |
| BMI age 34 | 25.9 | 4.7 | 25.2 | 4.1 |
| Education level age 34 | | | | |
| None | 2296 | 23.8% | 610 | 19.6% |
| Level 1 and below | 781 | 8.1% | 215 | 6.9% |
| O-levels | 2825 | 29.2% | 896 | 28.8% |
| A-levels | 623 | 6.4% | 215 | 6.9% |
| Degree/higher degree | 3140 | 32.5% | 1172 | 37.7% |
| Social class at age 42 | | | | |
| i | 511 | 6.2% | 221 | 7.2% |
| ii | 3688 | 44.4% | 1429 | 46.4% |
| iii non-manual | 1524 | 18.4% | 582 | 18.9% |
| iii manual | 1411 | 17.0% | 456 | 14.8% |
| iv partly skilled | 974 | 11.7% | 334 | 10.9% |
| v | 190 | 2.3% | 56 | 1.8% |
| Long standing illness/disability age 42 | | | | |
| No | 6918 | 70.9% | 2568 | 74.2% |
| Yes | 2845 | 29.1% | 892 | 25.8% |
| General health age 42 | | | | |

| | | | | |
|---|---|---|---|---|
| Poor | 437 | 4.5% | 81 | 2.3% |
| Fair | 1051 | 10.7% | 286 | 8.2% |
| Good | 6197 | 63.2% | 2208 | 63.7% |
| Excellent | 2114 | 21.6% | 892 | 25.7% |
| Marital status age 42 | | | | |
| used to be married/single | 3687 | 37.5% | 1169 | 33.7% |
| married/cohabiting | 6140 | 62.5% | 2297 | 66.3% |
| Main activity age 42 | | | | |
| Unemployed | 1451 | 14.8% | 363 | 10.5% |
| Employed | 8354 | 85.2% | 3099 | 89.5% |
| Education level age 42 | | | | |
| None | 2895 | 29.4% | 818 | 23.6% |
| Level 1 and below | 651 | 6.6% | 212 | 6.1% |
| O-levels | 2433 | 24.7% | 865 | 24.9% |
| A-levels | 540 | 5.5% | 213 | 6.1% |
| Degree/higher degree | 3322 | 33.8% | 1361 | 39.2% |
| Malaise score at age 42 | | | | |
| High malaise 4+ | 1578 | 18.4% | 473 | 15.1% |
| Low malaise 0-3 | 7000 | 81.6% | 2661 | 84.9% |
| BMI age 42 | 26.9 | 5.2 | 26.0 | 4.6 |
| **Exposure measures** | | | | |
| Member in uniformed organisations age 10-16 | | | | |
| No | 3412 | 54.7% | 975 | 50.6% |
| Yes | 2822 | 45.3% | 953 | 49.4% |
| Member in groups, clubs and associations age 30 | | | | |
| No | 8131 | 72.6% | 2258 | 69.2% |
| Yes | 3072 | 27.4% | 1003 | 30.8% |
| Member in groups, clubs and associations age 34 | | | | |
| No | 4878 | 50.7% | 1481 | 47.7% |
| Yes | 4752 | 49.3% | 1626 | 52.3% |
| Member in groups, clubs and associations age 42 | | | | |
| No | 3844 | 45.2% | 1233 | 39.5% |
| Yes | 4657 | 54.8% | 1889 | 60.5% |
| Social participation Index 16-42 | | | | |
| None | 4491 | 33.6% | 739 | 20.7% |
| Low | 4422 | 33.1% | 1084 | 30.3% |
| Medium | 2804 | 21.0% | 1016 | 28.4% |
| High | 1653 | 12.4% | 735 | 20.6% |
| **Outcome measures** | | | | |
| Mean activity time of MVPA (hr/d) | 0.9 | 0.4 | 0.9 | 0.4 |
| Mean daily step count | 9488.2 | 3691.6 | 9570.2 | 3482.2 |

**Supplemental Table 2: Description of potential confounding, exposure, and outcome variables**

| Variable | Age | Description/coding |
|---|---|---|
| Sex | 0 | A binary variable was used with information recorded at birth.<br><br>*(Men/ Women)* |
| Teen mother | 0 | The variable was derived using the age of the mother during CM's birth. Teenage mothers were classified those under the age of 20 years.<br><br>*(Teen pregnancy/ Non-teen pregnancy)* |
| Marital status of the mother | 0 and 16 | Information on the official marital status of the mother was recorded. At age 16, the participants were asked whether they lived with their natural parents now.<br><br>*(Married / Used to be married or single)* |
| Parental education | 0 | Information gathered on parental education reflected at which age mother and father completed education. A cut off point for minimum age for school leaving at the age of 16 was applied.<br><br>*(Left school before min age/ Stayed at school)* |
| Parental employment | 0 | Information on the employment status of mother and father.<br><br>*(Employed/ Unemployed)* |
| Birth weight | 0 | Recorded in grams and treated as a continuous variable. |

| | | |
|---|---|---|
| Smoking during pregnancy | 0 | Information on whether the mother was smoking during pregnancy and the number of cigarettes daily. A binary indicator was created.<br><br>*(Smoking/ Never or Used to smoke)* |
| Social class of father | 0 and 16 | Occupation of the father was coded according to the Registrar General's classification. Participant's current or most recent jobs were classified in 2 categories:<br><br>*Non-manual (i (professional), ii (managerial and technical)), iii (skilled non-manual) / Manual (iii(manual), IV (partly-skilled)/ and V (unskilled))* |
| Access to house amenities | 5 and 16 | A sum score of access to house amenities including bathroom, lavatory inside the house, hot water and kitchen was constructed. A binary indicator was created to indicate sole use of those amenities.<br><br>*(Shared use of amenities/ Sole use of amenities)* |
| House tenure | 5 | This variable indicates the tenure of the accommodation (owned outright, being bought, council rented, private rented but unfurnished, private rented but furnished, tied to occupation and other). A binary indicator was constructed to indicate the tenure.<br><br>*(Owner/ Other)* |
| Crowdedness in the house | 5 | Information on the number of persons per room. Crowdedness was classified in the 2 following categories:<br><br>*(Less than 2 people/ 2 people or more per room)* |

| | | |
|---|---|---|
| Family moves | 5 | Number of times the family has moved since the birth of the child. A binary variable was created.<br><br>(*0-2 times/ 3 or more times*) |
| Breastfeeding | 5 | The variable includes details on whether the CM was breastfed, as reported by the mother. A binary indicator was created.<br><br>*(Never breastfed/ Breastfed)* |
| Maternal mental health | 5 | A 24-tem Malaise Inventory was completed by the mothers of the CM. The variable was derived with a score of 8+ indicating depression.<br><br>*(Low malaise 0-7/ High malaise 8+)* |
| Bed wetting | 5 | Information provided by the mother on whether the child ever wets the bed at night.<br><br>*(Yes/ No)* |
| Cognitive ability | 10 | The cognitive ability of the CM was assessed using the British Ability Scales (BAS) that contains four sub-tests (word similarities and word definitions were used to capture verbal ability and recall of digits and matrices to capture non-verbal ability). (1)<br><br>In line with previous studies (2), a general cognitive ability factor was derived using Principal Component Analysis (PCA) for verbal and non-verbal ability scales. |

| | | |
|---|---|---|
| Health conditions | 10 | Parents were asked in a medical visit if the CM experienced the last 12 months a number of medical conditions (e.g. eczema, sore throats, ear infection, hay fever, heart condition, bronchitis, pneumonia, hearing loss, abdominal pain). A sum score of those conditions was derived.<br>A binary indicator was created with those having 3 or more/ less than 3 conditions:<br><br>*(Medical conditions 3 or more/ Medical conditions less than 3)* |
| Hospital admissions | 10 | The variable recorded the number of admissions in the hospital until the age of 10. |
| Body Mass Index (BMI) | 10, 16, 30, 34 and 42 | Trained medical personnel measured using standard protocols the height and weight of the CM. We used the harmonised measures of BMI ($kg/m^2$) by the CLOSER consortium. (3) |
| Externalising problems | 16 | Mothers of the participants completed the 19-item version of the Rutter behaviour questionnaire as part of the home interview. It is a measure of mental health capturing conduct problems, hyperactivity, emotional and peer problems.(4) Similarly to previous work (5), two scales were created, with 2 scales reflecting externalising and internalising.<br><br>Each item is scored in a 3-point response scale (“Not all true”=0, “Partly true”=1, “Certainly true”=2). Sum scores were created for each of the items of the internalising (e.g. often worried, bites nails) and externalising (e.g. destroys belongings, fights with others) scales.<br>Higher scores indicate higher internalising or externalising problems. Scores for the internalising scale may range from 0 to 16 and 0 to 22 for the externalising scale. |
| Internalising problems | 16 | |

| | | |
|---|---|---|
| Physical ability | 16 | Mothers reported if the CM has any disability, handicap, or impairment until this age.<br><br>*(Yes/ No)* |
| Social class | 30, 34 and 42 | Occupation of the CM was coded according to the Registrar General's classification. And then in 2 categories:<br><br>*Non-manual (i (professional), ii (managerial and technical)), iii (skilled non-manual) / Manual(iii(manual), IV (partly-skilled)/ and V (unskilled))* |
| Educational attainment | 30, 34 and 42 | Participants self-reported the highest level of qualification they obtained. The derived variable had two categories:<br><br>*(Below degree/ Degree or higher degree)* |
| Marital status | 30, 34 and 42 | CM self-reported their current, legal marital status. Participants in the "widowed" categories were included in the "Single or used to be married". When available at age 34 and 42 cohabiting participants were added in the married/cohabiting category.<br><br>*(Married or cohabiting/ Single or used to be married)* |
| Employment | 30, 34 and 42 | Information on the CM's current main activity:<br><br>*(Employed/ Unemployed)* |
| Mental health morbidity | | Malaise inventory was self-administrated to participants at different sweeps (4). At ages 16 and 30, a 24-item questionnaire was used. A shorter 9-item version was used for at age 34 and 42 (6). |

| | | |
|---|---|---|
| | 16, 30 | *(Low malaise 0-7/ High malaise 8+)* |
| | 34, 42 | *(Low malaise 0-3/ High malaise 4+)* |
| Long Standing Illness | 30, 34 and 42 | CM reported if they had any long standing and limiting illness, disability of infirmity likely to affect them for a period.<br><br>*(Yes/ No)* |
| Self-rated health | 30, 34 and 42 | CM self-reported on their own general health. A binary classification was created:<br><br>*(Poor or Fair/ Good or Excellent)* |
| **Exposure variables** | | |
| Social participation | 16 | CM were asked if they belonged to any uniformed youth organisations. A binary indicator of social participation was derived.<br>• Yes: belong/go to one<br>• No: used to go but not now<br>• No: never been/belonged)<br><br>*(Yes/No)* |
| Social participation | 30 | A binary social participation variable was derived by 2 variables to remain consistent with the other sweeps. CM were asked if they were a member of a Trade union or staff association:<br>• Yes - a trade union<br>• Yes - staff association<br>• Both<br>• No |

| | | |
|---|---|---|
| | | And whether they were members of any of those organisations:<br>• Political party<br>• Environmental group<br>• Other charity/voluntary groups?<br>• Women's group<br>• Women's Institute / Townswomen's Guild<br>• Parents'/School association<br>• Tenants'/Residents' group or neighbourhood watch<br>• None of these<br><br>*(Yes/No)* |
| Social participation | 34 | CM were asked if they have been involved with any groups or organisations from the following. A binary indicator was derived.<br>• Youth or children's activities, including school activities<br>• Politics, human rights, religious groups<br>• Environment, animal concerns<br>• Other voluntary or charity groups<br>• Local community or neighbourhood groups (including elderly, disabled, homeless)<br>• Hobbies, recreation, arts, social clubs<br>• Trade Union activity<br>• Other<br>• None of these<br><br>*(Yes/No)* |
| Social participation | 42 | CM were asked if they taking part in any of the following activities or groups. Similary to previous waves, a binary indicator was derived. |

| | | |
|---|---|---|
| | | • Political party<br>• Trade union<br>• Environmental group<br>• Parents'/School association<br>• Tenants'/Residents' group or neighbourhood watch<br>• Religious group or church organisation<br>• Voluntary service group<br>• Other community or civic group<br>• Social club/Working men's club<br>• Sports club<br>• Women's Institute / Townswomen's Guild<br>• Women's group / Feminist organisation<br>• Professional organisation<br>• Scouts/Guides organisation<br>• Any other organisation<br>• None of these<br><br>*(Yes/No)* |
| **Outcome variables** | | |
| Step count | 46 | Device-measured mean daily step count |
| Moderate to Vigorous physical activity (MVPA) | 46 | Device-measured mean activity time of moderate to vigorous intensity physical activity over the day (hr/day) |

### Supplemental text 1. Missing data strategy

Multiple Imputation with Chained Equations (MICE) was carried out using Stata 17.0 (7, 8). Multiple imputation works under the Missing at Random (MAR) assumption which is largely untestable. The MAR assumption implies that the observed data in a sample can explain the systematic differences between the missing and the observed values (9, 10).

To maximise the plausibility of the MAR assumption and restore sample representativeness, we used all the variables from the substantive analytical model and several auxiliary variables into the imputation model (11, 12). The auxiliary variables (physical activity at age 30 and 34 and smoking, mental health morbidity, long-standing illness, BMI, general self-rated health, housing tenure and marital status at age 46) that were not in the substantive analytical model were related with the missingness in the outcome variables (see Supplemental Table 3).

**Supplemental Table 3: Description of auxiliary variables**

| Variable | Age | Description |
|---|---|---|
| Physical activity | 30 and 34 | CM were asked if they do any regular (at least once per month for most of the year) exercise.<br><br>*(Yes/ No)* |
| Smoking behaviour | 46 | CM self-reported whether smokes (or used to smoke). A binary variable was created.<br><br>*(Never or ex-smoker/ Smoker)* |
| Mental health morbidity | 46 | Nine out of the original 24 self-completion questions which were combined to measure levels of psychological distress, or depression (13).<br><br>*(Low malaise 0-3/ High malaise 4+)* |
| Long Standing Illness | 46 | Any physical or mental health conditions or illnesses lasting or expected to last 12 months as reported by the CM.<br><br>*(Yes/ No)* |

| | | |
|---|---|---|
| Body Mass Index (BMI) | 46 | CMs had their height and wight measure by a nurse. Then a derived variable was calculated with the formula: Weight/((Height/100)*(Height/100)). |
| Self-rated health | 46 | CMs self-reported on their own general health. A binary classification was created.<br><br>(Poor or Fair/ Good or Excellent) |
| House tenure | 46 | As in previous sweeps a binary indicator was constructed to indicate the tenure.<br><br>*(Owner/ Other)* |
| Marital status | 46 | CM self-reported their current, legal marital status. Participants in the “widowed”, “divorced” or “former civil-partner” categories were included in the “Single or used to be married”.<br><br>*(Married or cohabiting/ Single or used to be married)* |

**Supplemental Table 4: Unadjusted regression coefficients (95% CIs) estimating the association between social participation and physical activity at age 46 (n = 3,646) for a. Accumulation and b. Sensitive Period Model**

| | | Mean daily step count (n=3,646) | Moderate to Vigorous Physical Activity (MVPA) (hr/day) (n=3,646) |
|---|---|---|---|
| a. | Accumulation Model: Social Participation Index[a] | | |
| | Low | 0.032(-0.003 - 0.066)* | 0.045(0.007 - 0.083)** |
| | Medium | 0.054(0.020 - 0.089)*** | 0.079(0.040 - 0.117)*** |
| | High | 0.071(0.033 - 0.109)*** | 0.092(0.051 - 0.134)*** |
| b. | Sensitive Period Model | | |
| | Age 16 | -0.002(-0.034 - 0.029) | -0.004(-0.038 - 0.030) |
| | Age 30 | 0.024(-0.003 - 0.051)* | 0.032(0.001 - 0.062)** |
| | Age 34 | 0.010(-0.016 - 0.037) | 0.016(-0.014 - 0.045) |
| | Age 42 | 0.071(0.045 - 0.098)*** | 0.078(0.049 - 0.107)*** |

*** p<0.01, ** p<0.05, * p<0.1

Notes: "None" was the reference category if cohort members had a negative response in all four sweeps (=0), “Low” if they engaged with activities only one time point (=1), “Medium” 2 times (=2) and “High” three times or more (≥3).

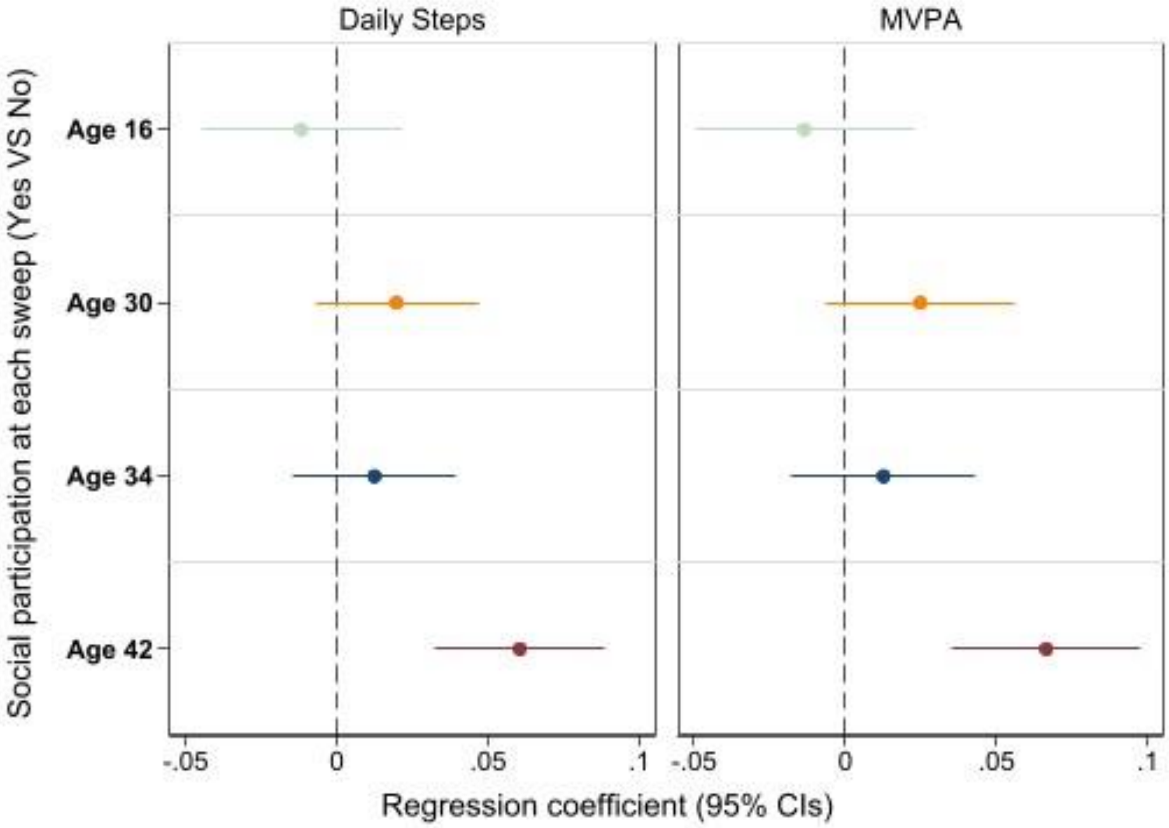


**Supplemental Figure 1: The figure represents adjusted regression coefficients (95% CI) for the b. Sensitive period Model by each physical activity measure**

**Supplemental Table 5: Adjusted regression coefficients (95% CIs) estimating the association between social participation and physical activity at age 46 (n = 3,646) for the Accumulation model with different sample restrictions**

| | Sample with participants with 7 days of accelerometer data (n=3,646) | | a. Sample with participants with 1 day of accelerometer data (n=5,569) | | b. Sample with participants at the Biomedical survey (n=8,581) | | c. Sample with participants not migrated and dead (n=17,237) | |
|---|---|---|---|---|---|---|---|---|
| | Mean daily Steps | MVPA | Mean daily Steps | MVPA | Mean daily Steps | MVPA | Mean daily Steps | MVPA |
| **Social Participation Index - Low** | 0.028 | 0.040** | 0.031** | 0.038** | 0.043*** | 0.049*** | 0.047*** | 0.053*** |
| | (-0.006 - 0.063) | (0.002 - 0.079) | (0.001 - 0.061) | (0.006 - 0.070) | (0.012 - 0.074) | (0.017 - 0.081) | (0.016 - 0.078) | (0.020 - 0.086) |
| | 0.109 | 0.039 | 0.042 | 0.020 | 0.006 | 0.003 | 0.003 | 0.002 |
| **Social Participation Index - Medium** | 0.048*** | 0.068*** | 0.065*** | 0.075*** | 0.072*** | 0.082*** | 0.074*** | 0.085*** |
| | (0.013 - 0.084) | (0.029 - 0.108) | (0.033 - 0.096) | (0.042 - 0.109) | (0.040 - 0.104) | (0.048 - 0.116) | (0.035 - 0.112) | (0.045 - 0.125) |
| | 0.008 | 0.001 | 0.000 | 0.000 | 0.000 | 0.000 | 0.000 | 0.000 |
| **Social Participation Index - High** | 0.059*** | 0.073*** | 0.075*** | 0.079*** | 0.081*** | 0.084*** | 0.083*** | 0.087*** |
| | (0.019 - 0.098) | (0.029 - 0.117) | (0.040 - 0.110) | (0.041 - 0.117) | (0.044 - 0.118) | (0.045 - 0.123) | (0.038 - 0.129) | (0.042 - 0.132) |

The reference group for all models is "Never participated". *** p<0.001, ** p<0.01, * p<0.05.
Notes: Models adjusted for confounders until age 16.

**Supplemental Table 6: Adjusted regression coefficients (95% CIs) estimating the association between social participation (sports clubs omitted) and physical activity at age 46 (n = 3,646) for a. Accumulation and b. Sensitive Period Model**

| | Mean daily step count (n=3,646) | Moderate to Vigorous Physical Activity (MVPA) (hr/day) (n=3,646) |
|---|---|---|
| **a. Accumulation Model: Social Participation Index[a]** | | |
| **Low** | 0.017(-0.017 - 0.051) | 0.020(-0.017 - 0.057) |
| **Medium** | 0.040(0.005 - 0.075)** | 0.057(0.017 - 0.096)*** |
| **High** | 0.046(0.007 - 0.085)** | 0.054(0.010 - 0.097)** |
| **b. Sensitive Period Model [a]** | | |
| **Age 16** | -0.011(-0.044 - 0.021) | -0.013(-0.049 - 0.023) |
| **Age 30** | 0.020(-0.007 - 0.047) | 0.025(-0.006 - 0.056) |
| **Age 34** | 0.013(-0.014 - 0.039) | 0.013(-0.018 - 0.043) |
| **Age 42** | 0.043(0.016 - 0.069)*** | 0.045(0.015 - 0.074)*** |

*** p<0.01, ** p<0.05, * p<0.1

Notes: "None" was the reference category if cohort members had a negative response in all four sweeps (=0), "Low" if they engaged with activities only one time point (=1), "Medium" 2 times (=2) and "High" three times or more (≥3)
[a]All models adjusted as in the main analysis

**Supplemental Table 7: Adjusted regression coefficients (95% CIs) estimating the association between social participation (age 30 omitted) and physical activity at age 46 (n = 3,646) for a. Accumulation and b. Sensitive Period Model**

| | Mean daily step count (n=3,646) | Moderate to Vigorous Physical Activity (MVPA) (hr/day) (n=3,646) |
|---|---|---|
| **a. Accumulation Model: Social Participation Index[a]** | | |
| **Low** | 0.016(-0.016 - 0.049) | 0.036(0.000 - 0.072)** |
| **Medium** | 0.042(0.006 - 0.077)** | 0.060(0.021 - 0.099)*** |
| **High** | 0.050(0.001 - 0.099)** | 0.060(0.006 - 0.115)** |
| **b. Sensitive Period Model [a]** | | |
| **Age 16** | -0.013(-0.045 - 0.020) | -0.013(-0.051 - 0.025) |
| **Age 34** | 0.012(-0.014 - 0.037) | 0.012(-0.017 - 0.042) |
| **Age 42** | 0.059(0.032 - 0.087)*** | 0.066(0.035 - 0.096)*** |

*** p<0.01, ** p<0.05, * p<0.1

Notes: "None" was the reference category if cohort members had a negative response in all four sweeps (=0), "Low" if they engaged with activities only one time point (=1), "Medium" 2 times (=2) and "High" three times (=3)

[a]All models adjusted as in the main analysis
